# Recent-Onset and Persistent Tinnitus: Uncover the Differences in Brain Activities using Resting-State Functional Magnetic Resonance Imaging Technologies

**DOI:** 10.1101/2022.06.26.22276922

**Authors:** Haoliang Du, Xu Feng, Xiaoyun Qian, Jian Zhang, Bin Liu, Ao Li, Zhichun Huang, Xia Gao

## Abstract

**Objective:** This project aimed to investigate the differences in the intra-regional brain activity and inter-regional functional connectivity between patients with recent-onset tinnitus and persistent tinnitus using resting-state functional magnetic resonance imaging (rs-fMRI) technologies, including the Amplitude of Low-Frequency Fluctuations (ALFF), regional homogeneity (ReHo), and Voxel-Wise Functional Connectivity (FC).

**Method:** We acquired rs-fMRI scans from 82 subjects (25 subjects without recent-onset tinnitus, 28 subjects with persistent tinnitus, and 29 subjects as healthy control). Age, gender, and year of education were matched across all three groups. We performed ALFF, ReHo, and Voxel-Wise Functional Connectivity (FC) for all subjects.

**Result:** Compared with the control group (CN), subjects with recent-onset tinnitus (ROT) and with persistent tinnitus (PT) manifested significantly reduced ALFF and ReHo activity within the left and right dorsolateral superior frontal gyrus (SFG) and Gyrus Rectus (GR). Additional Voxel-Wise Functional Connectivity (FC) revealed decreased connectivity between the dorsolateral SFG (left and right) and right Superior Parietal Gyrus (SPG), right Middle Frontal Gyrus (MFG), and left medial Superior Frontal Gyrus (mSFG) within these two groups. Significant differences were observed between the ROT and PT groups, with the ROT group demonstrating reduced functional connectivities.

**Conclusion:** Upon analyzing our data, we suggested that patients with persistent tinnitus have more difficulty monitoring external stimuli and reorienting attention than patients with recent-onset tinnitus. In addition, patients who perceive higher levels of disruption from tinnitus are more likely to develop persistent and debilitating tinnitus once the tinnitus lasts longer than six months. Therefore, we strongly recommend that clinicians implement effective tinnitus management strategies for patients with recent-onset tinnitus as soon as possible.

## 1 Introduction

Subjective tinnitus is a conscious auditory perception without a corresponding external source. Subjective tinnitus represents one of the most common yet distressing otologic pathologies, affecting approximately 8 to 20% of the adult population (Roberts et al., 2010). Existing studies reported that subjective tinnitus is commonly associated with hearing loss, cerumen impaction, middle and inner ear-related pathologies, noise exposure, exposure to ototoxic medications and chemicals, aging, insomnia, anxiety, depression, head and neck injuries, and temporomandibular joint (TMJ) dysfunction (Baguley et al., 2013; Tunkel et al., 2014; Makar, 2021). In addition, tinnitus can be persistent, bothersome, and costly for patients and societies in general. Cases of patients with extraordinarily persistent and debilitating tinnitus accompanied by severe anxiety or depression attempting suicide have been reported (Szibor, Mäkitie and Aarnisalo, 2019).

It is generally believed that lesions in the peripheral hearing system and neuronal changes within the central nervous system contribute to tinnitus. Kapolowicz and Thompson reported that tinnitus might be closely related to an imbalance between the auditory neuronal excitation and inhibition network, leading to plasticity changes in the central auditory system (Kapolowicz & Thompson, 2020). Knipper et al. (2021) proposed that hearing loss may contribute to a top-down mechanism that leads to tinnitus perception (Knipper et al., 2021). Khan and colleagues suggested that tinnitus might be a compensatory response to damage to the peripheral hearing system (Khan et al., 2021). Cai, Xie, and colleagues reported abnormal functional connectivity in the auditory and non-auditory cortices in patients with hearing loss and tinnitus (Cai et al., 2020). Zhou and colleagues suggested that patients with hearing loss and tinnitus demonstrated abnormal intra-regional neural activity and disrupted connectivity in the hub regions of some non-auditory networks, including the default mode network (DMN), optical network, dorsal and ventral attention network (DAN & VAN), and central executive network (CEN) (Zhou et al., 2019). Minami and colleagues reported that tinnitus patients with hearing loss showed a statistically significant reduction in auditory-related functional connectivity than the control group (Minami et al., 2018). Finally, our previous project, using ALFF, ReHo, and Voxel-Wise Functional Connectivity technologies, revealed that disruptions in brain regions responsible for attention and stimuli monitoring and orientations could lead to tinnitus (Du et al., 2022).

Tinnitus has different forms, degrees of severity, and onset duration, which can only be described by patients’ testimony and corresponding symptoms. When categorizing tinnitus based on its duration of onset (recent-onset or persistent), numerous studies have concentrated on developing pathophysiological models for chronic tinnitus (tinnitus that has a duration of onset of at least six months). However, few have investigated the neuronal changes that occur from recent-onset to persistent tinnitus (Stolzberg et al., 2013; Cai et al., 2020; Lan et al., 2020). To our best knowledge, we have not located any studies investigating this issue using resting-state functional magnetic resonance imaging (rs-fMRI) technologies. Furthermore, investigating this issue will be critical for identifying contributing neural mechanisms and possible interventions to stop this transition. Therefore, our project aims to uncover the differences in brain activity using Resting-State Functional Magnetic Resonance Imaging (rs-fMRI) technologies between recent-onset tinnitus patients and persistent tinnitus patients and apply our findings to existing tinnitus management strategies.

## 2 Method

### 2.1 Subjects’ Demographic and Clinical Information

The Research Ethics Committee of the Affiliated Zhongda Hospital of Southeast University approved this study. All individuals provided written informed consent before they participated in the study. We recruited eighty-two subjects (all right-handed, with at least eight years of education), including twenty-five tinnitus subjects with recent-onset tinnitus (ROT), twenty-eight tinnitus subjects with persistent tinnitus (PT), and twenty-nine healthy subjects as the control group (CN) through our outpatient clinics between September 2011 and September 2013. The patients were group-matched in terms of age, sex and education. Twenty-five subjects perceived bilateral tinnitus, and the rest, twenty-eight subjects, perceived unilateral tinnitus. We defined the time course of tinnitus (recent-onset or persistent) according to the Tinnitus Clinical Practice Guideline from the American Academy of Otolaryngology-Head and Neck Surgery. According to the guideline, if the overall duration of onset equals or is less than six months, the tinnitus will be determined to be recent-onset. If the overall duration of onset is more than six months, the tinnitus will be defined as persistent (Tunkel et al., 2014).

We performed pure-tone audiometric testing (PTA for 250, 500, 1000, 2000, 4000, 6000, and 8000 Hz) for all recruited subjects. Subjects with 7-frequency PTA below 25 dB HL were considered clinically normal hearing. In addition, we performed comprehensive tympanometry, diagnostic distortion-product otoacoustic emissions (DPOAE), and diagnostic auditory brainstem response (ABR) for all subjects to rule out middle ear pathologies and auditory neuropathy (ANSD). Furthermore, we collected crucial information about the duration of tinnitus and the presence of insomnia from all subjects.

To assess the severity and distress associated with tinnitus, we distributed the Iowa version of the tinnitus handicap questionnaire (THQ) (Kuk et al., 1990) to both the ROT and PT groups. We also distributed the Self-Rating Depression Scale (SDS) and the Self-Rating Anxiety Scale (SAS) questionnaires to all subjects for anxiety and depression screening (Zung, 1986; Zung, 1971). No significant group differences were discovered for subjects’ gender, age, and educational background (p > 0.05). However, we did discover a statistically significant difference in subjects’ THQ total score, SAS, and SDS scores between groups (p < 0.05). Subjects’ demographic and clinical characteristics for each group were summarized in table 1.

**Table 1.**
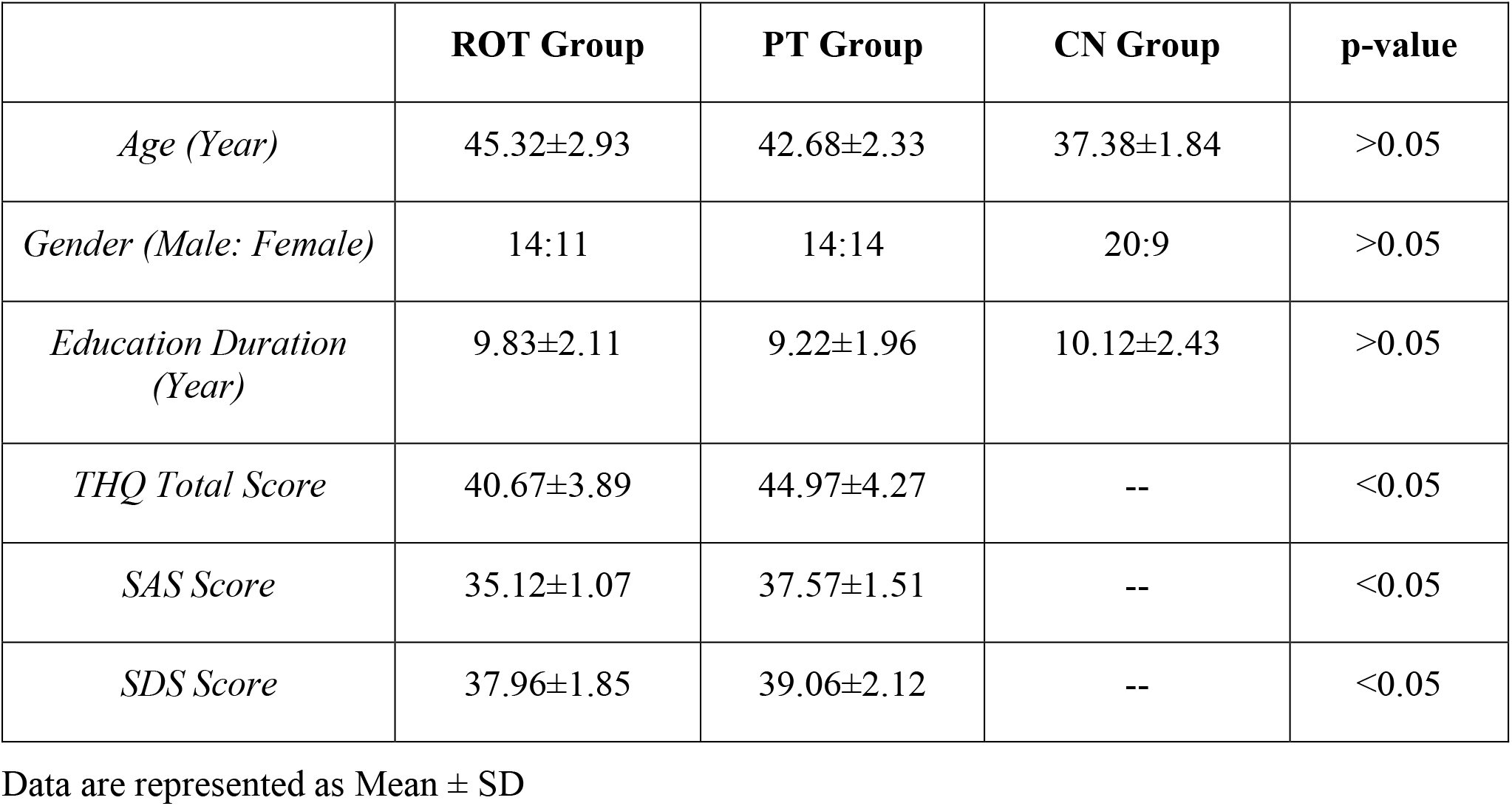
Subject Characteristics of the Recent-onset Tinnitus Group (ROT), Persistent Tinnitus Group (PT), and Control Group (CN)

### 2.2 Subject Exclusion Criteria

The exclusion criteria for this study included Meniere’s disease, objective tinnitus, pulsatile tinnitus, histories of consuming alcohol, severe smoking, head and neck injuries, epilepsy, stroke, Alzheimer’s disease, Parkinson’s disease, cancer, MRI contraindications, primary psychiatric conditions including Generalized Anxiety Disorder (GAD), depression and Schizophrenia, and severe visual impairment. None of our subjects failed the depression and anxiety screening.

### 2.3 fMRI Scanning & Data Acquisition

We acquired the imaging data using a 3.0 T MRI scanner (Siemens MAGENETOM Trio, Erlangen, Germany) with a standard head coil. We provided all subjects with foam paddings and earmuffs to minimize head motion and noise exposure during the scanning process. The subjects were instructed to remain calm during the scan with their eyes closed without falling asleep or thinking of anything particular. Functional images were obtained axially using a gradient echo-planar sequence sensitive to BOLD contrast as follows: repetition time (TR) =2000 ms; echo time (TE) = 25 ms; slices = 36; thickness = 4 mm; gap = 0 mm; field of view (FOV) = 240 × 240 mm; acquisition matrix = 64 × 64; and flip angle (FA) = 90°.

### 2.4 Amplitude of Low-Frequency Fluctuations (ALFFs): Preprocessing & Analysis

Resting-state ALFF can reflect spontaneous neural activity and yield physiologically meaningful results. Pre-processing of the ALFF images was performed using the toolbox Data Processing Assistant for Resting-State fMRI (DPARSF 5.2), Statistical Parametric Mapping (SPM 12), and Matlab 2021b. We removed the first five volumes from each time series to account for subjects’ adaptation to the scanning environment. Slice timing and re-alignment for head-motion correction were performed for the remaining 175 images. Afterward, we performed the following procedures: spatially normalized into the stereotactic space of the Montreal Neurological Institute (MNI) (resampling voxel size = 3 × 3 × 3 mm^3^) and smoothed using a Gaussian kernel of 6 mm full width at half-maximum (FWHM), de-trending, and filtering (0.01–0.08 Hz). The subjects with a head motion with more than 2.0 mm displacement or a 2.0-degree rotation in the *x, y*, or *z* directions were excluded from this study.

We then analyzed the ALFF data by transforming time to the frequency domain using Fast Fourier Transform. Next, we computed the square root of the power spectrum and averaged squared across 0.01–0.08 Hz at each voxel. The calculated averaged square root was taken as the ALFF. Finally, the ALFF of each voxel was divided by the global mean ALFF value for standardization (Du et al., 2022).

### 2.5 Regional Homogeneity (ReHo): Preprocessing & Analysis

The ReHo calculates the synchronization of low-frequency fluctuations between a given voxel and neighboring voxels, reflecting the neural function synchronization in the local brain region. Pre-processing of ReHo images was performed using the toolbox Data Processing Assistant for Resting-State fMRI (DPARSF 5.2), Statistical Parametric Mapping (SPM 12), and Matlab 2021b. We removed the first five volumes from each time series to account for subjects’ adaptation to the scanning environment. Slice timing and re-alignment for head-motion correction were performed for the remaining 175 images. The following procedures were performed: spatially normalized into the stereotactic space of the Montreal Neurological Institute (MNI) (resampling voxel size = 3 × 3 × 3 mm^3^), de-trending, and filtering (0.01–0.08 Hz).

After the pre-processing stage, we performed the image calculation using the Kendall coefficient of concordance of the time series of a given voxel with its 27 nearest neighbors. Next, ReHo analyses were calculated using the DPARSF 5.2 software. The ReHo value of each voxel was then standardized by partitioning the primal value using the global mean ReHo value. Finally, the data were smoothed with a Gaussian kernel of 6 mm full-width at half maximum (FWHM) for further statistical analysis (Du et al., 2022).

### 2.6 Voxel-Wise Functional Connectivity (FC): Pre-processing & Analysis

We performed the Voxel-Wise FC analysis using the toolbox Data Processing Assistant for Resting-State fMRI (DPARSF 5.2), Statistical Parametric Mapping (SPM 12), and Matlab 2021b. The first ten volumes were removed from each time series to account for subjects’ time to adapt to the scanning environment. Then, slice timing and re-alignment for head-motion correction were performed for the remaining 170 images. Afterward, the procedures were carried out as follows: spatially normalized into the stereotactic space of the Montreal Neurological Institute (MNI) (resampling voxel size = 3 × 3 ×3 mm^3^) and smoothed using a Gaussian kernel of 6 mm full width at half-maximum (FWHM), de-trending, and filtering (0.01–0.08 Hz). Subjects with a head motion with more than 2.0 mm displacement or a 2.0-degree rotation in the *x, y*, or *z* directions were excluded (Du et al., 2022).

We extracted the ALFF and ReHo differences in brain regions between recent-onset tinnitus subjects and persistent tinnitus subjects for Voxel-Wise FC analysis and defined them as seeds. We then used the average time series of seeds as a reference and calculated the Pearson correlation coefficient between the average signal change of each seed and the time sequences of other voxels in the brain. Finally, we converted the correlation coefficient to a z-value using Fisher’s z-transformation.

### 2.7 Statistical Analysis and Graphic Illustration

The one-way analysis of variance (ANOVA) was firstly conducted to test mean differences in ALFF, ReHo, and functional connectivity (FC) between the control group (CN), the group with recent-onset tinnitus (ROT), and the group with persistent tinnitus (PT) (Matlab 2021b). The statistically-significant difference between the groups was determined at p < 0.05. Subjects’ age and gender were included as nuisance covariates. Next, we applied Family-Wise Error (FWE) correction for multiple comparisons, using voxel-level inference at p < 0.001 and cluster-level inference at p<0.05. Two-sample t-tests were then conducted to investigate the ALFF, ReHo, and functional connectivity (FC) differences between subjects with recent-onset tinnitus (ROT) and control group (CN), subjects with persistent tinnitus (PT) and control group (CN), and subjects with recent-onset tinnitus (ROT) and subjects with persistent tinnitus (PT). Again, the statistically-significant difference between the groups was determined at p < 0.05. Finally, we used the MRIcroGL software to draw 2-dimensional brain images to display the brain areas with statistically significant differences.

## 3. Results

### 3.1 ALFF Results

We discovered significant ALFF value differences in the left and right dorsolateral SFG and left gyrus rectus (GR) for the ROT and PT groups compared to the CN group (Fig. 1). Compared with the control group (CN), ALFF’s T-value for both the ROT group and PT group in the left Gyrus Rectus (GR) were significantly lower than the global mean values from the control (CN) group (p < 0.05). The ALFF’s T-value for the ROT group is lower than the PT group (p < 0.05).

No statistical significance was discovered for the left dorsolateral Superior Frontal Gyrus (SFG) between the ROT and the PT groups (p > 0.05). However, compared with the control group (CN), ALFF’s T-value for both the ROT group and PT group in the right dorsolateral Superior-Frontal Gyrus (SFG) were significantly lower than the global mean values from the control (CN) group (p < 0.05).

No statistical significance was discovered for the right Gyrus Rectus (GR) between the ROT and the PT groups (p > 0.05). However, compared with the control group (CN), ALFF’s T-value for both the ROT group and PT group in the left Gyrus Rectus (GR) were significantly lower than the global mean values from the control (CN) group (p < 0.05). These results are demonstrated in table 2a.

**Table 2a.**
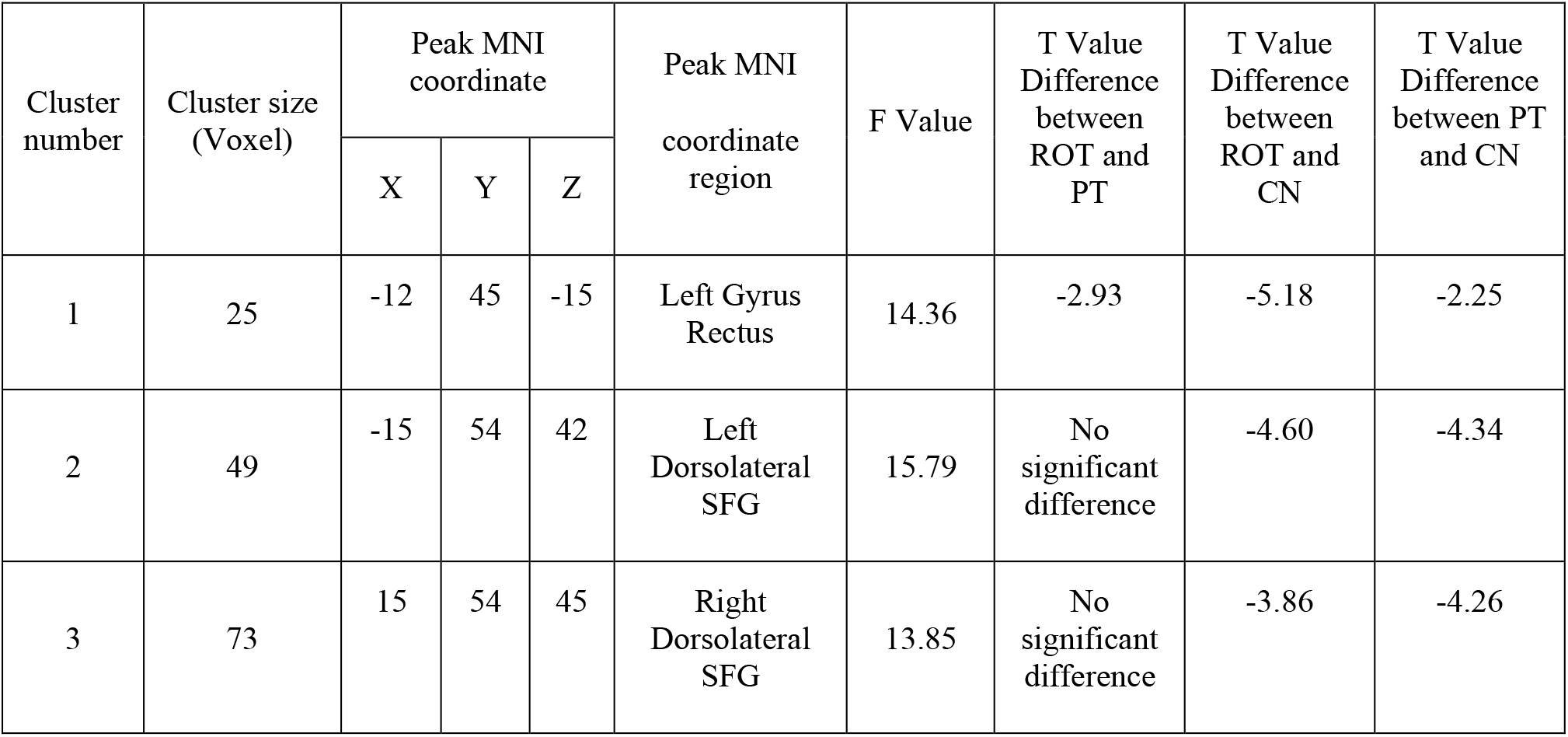
Decreased ALFF activities in both Recent-onset Tinnitus (ROT) and Persistent Tinnitus (PT) with than in the control group (CN)

### 3.2 ReHo Results

We also discovered significant ReHo value differences in the right dorsolateral SFG for both ROT and PT groups compared to the control (CN) group (Fig. 2). Regarding ReHo’s T-value, both the ROT group and PT group in the right dorsolateral SFG revealed significantly lower values than the global mean values from the control (CN) group (p < 0.05) (Table. 2b). A two-sample t-test did not reveal any statistical differences between the ROT group and the PT group in the right Dorsolateral Superior Frontal Gyrus (SFG) (p > 0.05).

**Table 2b.**
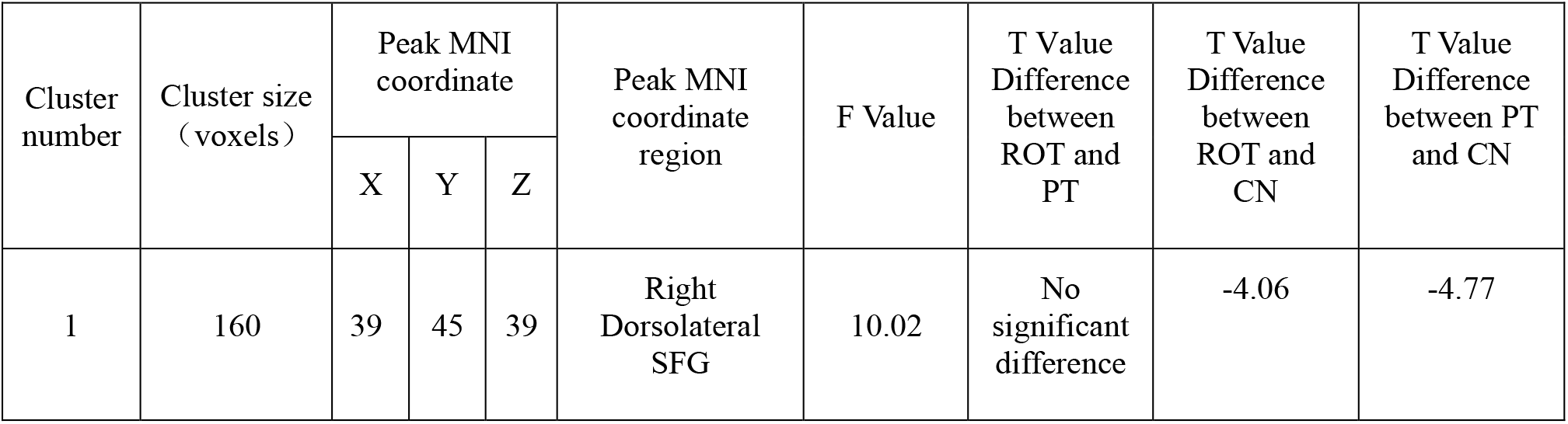
Decreased ReHo activities in both Recent-onset Tinnitus (ROT) and Persistent Tinnitus (PT) with than in the control group (CN)

### 3.3 Voxel-Wise Functional Connectivity (FC) Results

Two regions identified from the ALFF analysis (dorsolateral SFG, left and right) were used as seeds for further FC analysis. Brain regions with significant functional connectivity pattern differences for the ALFF analysis clusters 2 and 3 were demonstrated in Figures 3 and 4, respectively. In contrast to the control group (CN), both the ROT group and PT group exhibited a reduction in connectivity between the seed region in the left dorsolateral SFG (ALFF cluster 2) and right superior parietal gyrus (SPG), right dorsolateral superior frontal gyrus (SFG), and left medial superior frontal gyrus (SFG) (p < 0.05) (Table 3a). No difference was observed between the ROT and PT groups (p > 0.05). At the same time, both the ROT group and PT group exhibited decreased connectivity between the seed region in the right dorsolateral SFG (ALFF cluster 3) and right Middle Frontal Gyrus (MFG), left medial Superior Frontal Gyrus (SFG), and right Superior Parietal Gyrus (SPG) (p < 0.05) (Table 3b). No difference was observed between the ROT and PT groups (p > 0.05), except for a reduced connectivity pattern between the right dorsolateral SFG (ALFF cluster 3) and right Middle Frontal Gyrus (MFG) in the PT group than in the ROT group.

**Table 3a.**
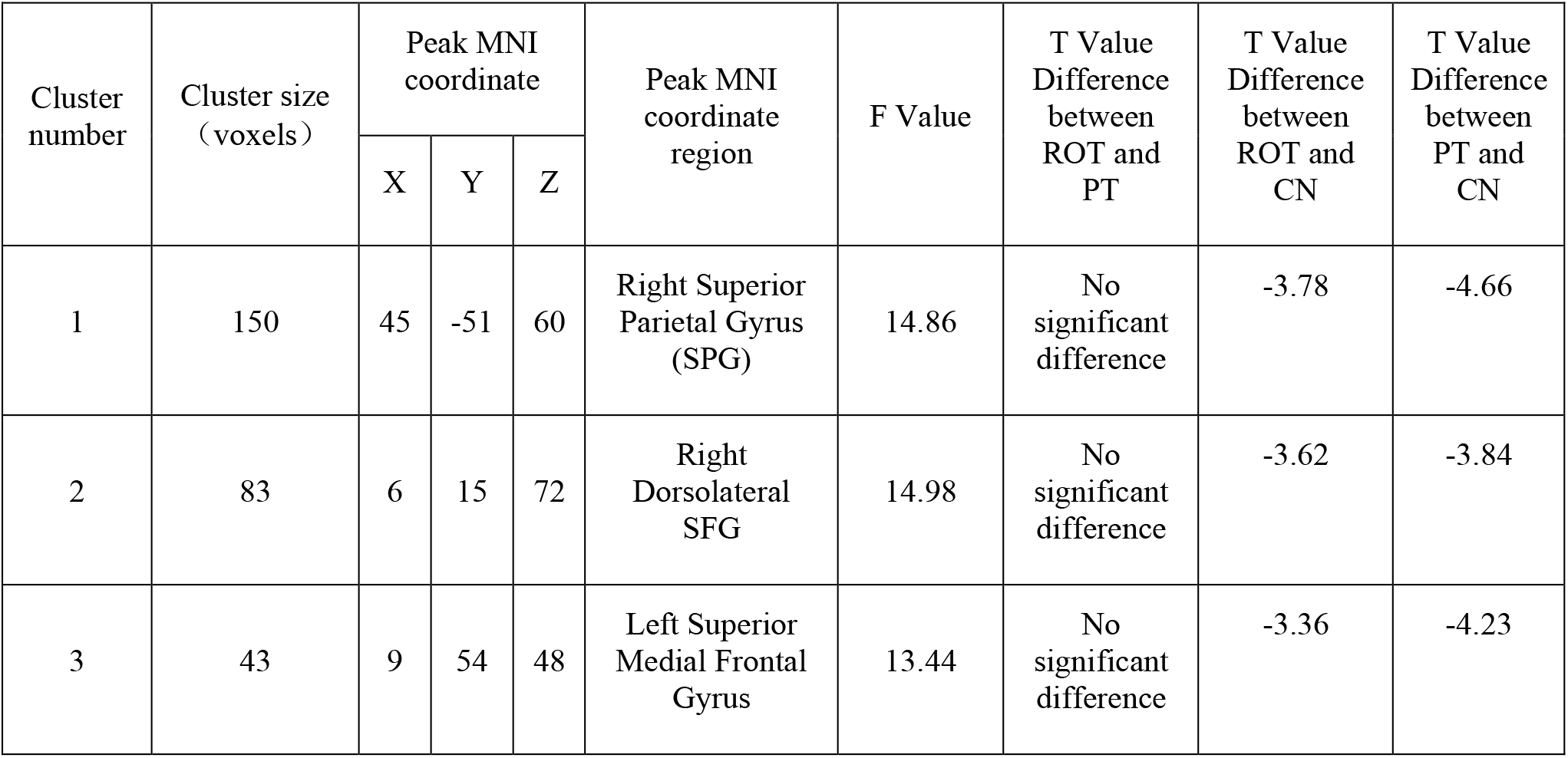
Decreased activities in Voxel-Wise Functional Connectivity (FC) ALFF cluster 2 for both Recent-onset Tinnitus (ROT) and Persistent Tinnitus (PT) groups than in the control group (CN)

**Table 3b.**
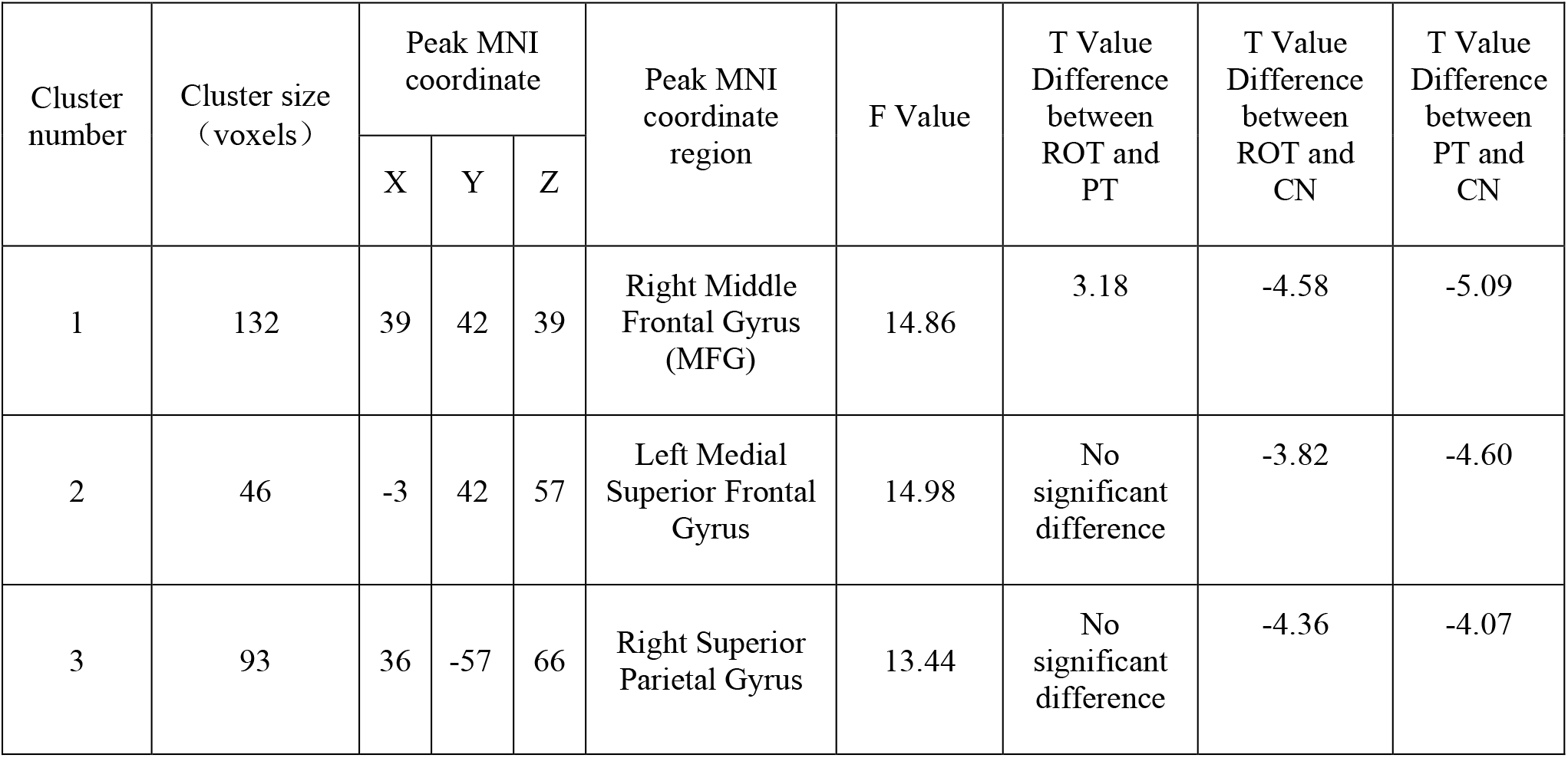
Decreased activities in Voxel-Wise Functional Connectivity (FC) ALFF cluster 3 for both Recent-onset Tinnitus (ROT) and Persistent Tinnitus (PT) groups than in the control group (CN)

One region identified from the ReHo analysis (right dorsolateral SFG) was used as seeds for further FC analysis. Brain regions with significant functional connectivity pattern differences were illustrated in Figure 5. In contrast to the control group (CN), both the ROT group and PT group demonstrated lower connectivity levels between the seed region in the right dorsolateral SFG and right middle frontal gyrus (MFG), left medial superior frontal gyrus, and right superior parietal gyrus (p < 0.05) (Table 3c). No difference was observed between the ROT and PT groups (p > 0.05), except for an elevated connectivity pattern between the right dorsolateral SFG (ReHo Cluster 1) and right Middle Frontal Gyrus (MFG) in the ROT group than in the PT group.

**Table 3c.**
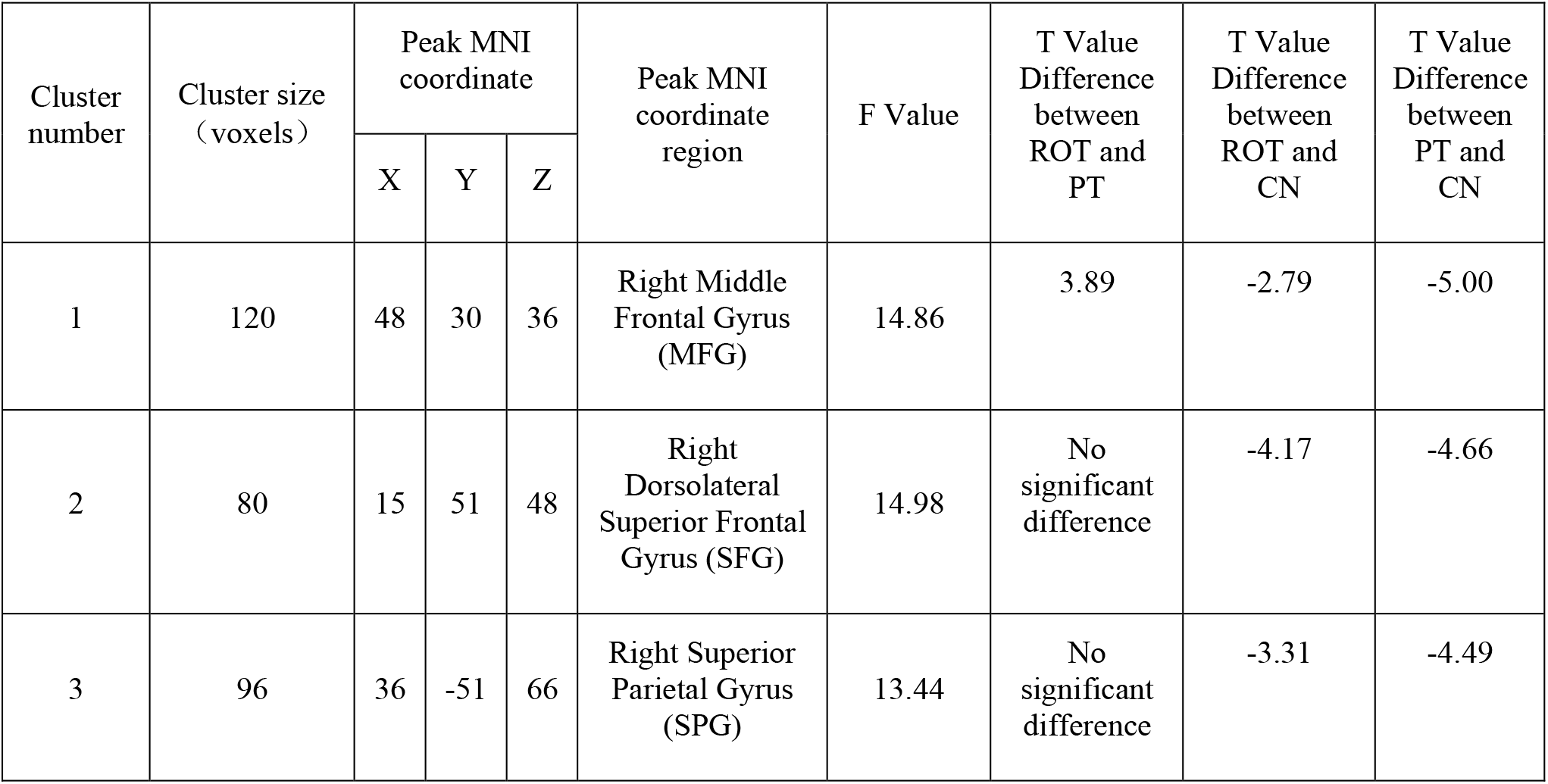
Decreased activities in Voxel-Wise Functional Connectivity (ReHo Cluster 1) for both Recent-onset Tinnitus (ROT) and Persistent Tinnitus (PT) groups than in the control group (CN)

## 4. Discussion

In the current study, we utilized various resting-state fMRI technologies, including the ALFF, ReHo, and Voxel-Wise functional connectivity (FC), to investigate the differences in the intra-regional brain activity and inter-regional functional connectivity in patients with recent-onset tinnitus (ROT) and persistent tinnitus (PT). To our best knowledge, this is the first study to reveal neuronal changes during the transition from recent-onset to persistent tinnitus using the resting-state fMRI technologies.

Our findings revealed that subjects with recent-onset and persistent tinnitus demonstrated abnormal intra-regional neural activity and disrupted functional connectivity. In addition, regions of some non-auditory networks involving the default mode network (DMN), optical network, dorsal attention network (DAN), and central executive network (CEN) were affected (Chen et al., 2017). Furthermore, we discovered significant differences within the ALFF, ReHo, and FC activity levels between the ROT and PT group, with the PT group demonstrating the lowest activity and connectivity level among all three groups. In order to identify the differences in brain activities between the recent-onset and persistent tinnitus subjects, we will explore the roles of each brain region revealed by the rs-FMRI analysis and identify possible strategies to prevent the transition from recent-onset tinnitus to persistent tinnitus.

### 4.1 Elevated Activity in Left Gyrus Rectus (GR) for Persistent Tinnitus Patients

The gyrus rectus (GR) is located at the most medial margin of the inferior surface of the frontal lobe. Although its specific function remains unclear, clinical reports indicated that patients who received surgical removal of the gyrus rectus demonstrated temporary cognitive deficits, including a reduction in memory and personality changes (Joo et al., 2016). In addition, studies using resting-state functional connectivity technologies revealed that patients with distressful tinnitus demonstrated abnormal brain activities within the bilateral gyrus rectus (Ueyama et al., 2013; Ueyama et al., 2015). Furthermore, studies also revealed that the gyrus rectus demonstrated anatomical connections with the limbic system (Lan et al., 2022). Du and colleagues reported that the gyrus rectus demonstrated strong functional connectivity with the anterior, medial and posterior orbital gyrus, superior frontal gyrus, ventromedial prefrontal cortex, and the anterior cingulate cortex (Du et al., 2020).

Our findings revealed that subjects with recent-onset tinnitus (ROT) demonstrated reduced activity levels at the gyrus rectus compared to subjects with persistent tinnitus (PT). In addition, compared to the healthy control group (CN), subjects from both tinnitus groups demonstrated reduced activity levels at the gyrus rectus. Therefore, these results indicate that patients with recent-onset or persistent tinnitus might perceive a temporary cognitive decline due to disruptions at the gyrus rectus. Furthermore, for patients with recent-onset tinnitus (ROT), the level of disruption to cognitive processing from tinnitus might be higher than those with persistent tinnitus due to the novelty of tinnitus.

### 4.2 Reduced Activity in Dorsolateral Superior Frontal Gyrus (SFG) for Both Recent-onset and Persistent Tinnitus Patient

Both ALFF and ReHo analysis revealed a reduction in activity level at dorsolateral SFG on both sides for subjects with persistent or recent-onset tinnitus. Results did not reveal significant differences in dorsolateral SFG activities between the recent-onset tinnitus group (ROT) and the persistent tinnitus group (PT). The main functions of the dorsolateral SFG comprise top-down processing and cognitive functions, including working memory, episodic memory, goal-driven attention, planning, problem-solving, and task-switching. These findings imply dorsolateral SFG’s role in the CEN manipulations (Kinoshita et al., 2012; Hu et al., 2016).

In addition, the dorsolateral SFG demonstrates functional connectivity with the Default Mode Network (DMN), especially the precuneus. Existing literature indicated that the DMN specialized in internally-oriented cognitive processes such as conceptual processing, daydreaming, and future planning (Cloutman & Lambon Ralph, 2012; Lin et al., 2017). Therefore, we suggest that the dorsolateral SFG regulates the interaction between the CEN and DMN. Reduced dorsolateral SFG activity might disrupt the CEN, eventually reducing patients’ top-down attention-filtering capability. Furthermore, our results suggested that the overall duration of tinnitus does not contribute to reduced activity levels at left and right dorsolateral SFG. Tinnitus patients can perceive difficulties switching their attention away from the tinnitus, regardless of experiencing recent-onset or persistent tinnitus.

### 4.3 Reduced Functional Connectivity between Bilateral Dorsolateral Superior Frontal Gyrus (SFG) and Medial Superior Frontal Gyrus (mSFG) in Both Recent-Onset and Persistent Tinnitus Subjects

Existing literature revealed that the medial SFG has anatomic connections with the cingulate cortex (mainly the anterior and medial section of the cingulate cortex, ACC & MCC) through the cingulum and that functional correlation with the MCC and the DMN (Nagahama et al., 1999). In addition, dense connections between the dorsolateral prefrontal cortex (DLPFC) (including the SFG) and the ACC and MCC have also been discovered in humans (Zhang et al., 2011; Cloutman & Lambon Ralph, 2012; Ueyama et al., 2013).

Moreover, the rs-FCs between the SFG, ACC, and MCC have been reported (Cloutman & Lambon Ralph, 2012; Yang et al., 2014; Khan et al., 2021). The anatomical and functional connections between the medial SFG and the anterior MCC suggest that the medial SFG is involved in cognitive control because the anterior part of the MCC has been related to cognitive control, including conflict monitoring, response selection, error detection, and attention manipulation. Additionally, the medial SFG demonstrates anatomic connections with the ACC, a core node of the DMN, and functional correlation with the DMN, suggesting that the medial SFG is critical for the DMN manipulation (Hu et al., 2021).

Our finding suggested that the overall duration of tinnitus onset does not play a role in generating functional connectivity differences within the left medial superior frontal gyrus. Nevertheless, subjects from the recent-onset (ROT) and persistent (PT) tinnitus group demonstrated reduced functional connectivity between the bilateral dorsolateral SFG and left medial superior frontal gyrus compared to the healthy control group (CN). As a result, we propose that reduced functional connectivity between the dorsolateral SFG and the medial SFG disrupts DMN regulation, further reducing patients’ ability to manipulate attention. Furthermore, this significant change within the top-down attention-regulating mechanism increases tinnitus perception, regardless of the overall duration of tinnitus onset.

### 4.4 Functional Connectivity Abnormality between Bilateral Dorsolateral Superior Frontal Gyrus (SFG) and Right Middle Frontal Gyrus (MFG) in Both Recent-Onset and Persistent Tinnitus Subjects

As a critical component of the ventral attention network (VAN), the right middle frontal gyrus (MFG) served as a convergence center for the DAN and the VAN by working as a circuit-breaker to interrupt ongoing endogenous attentional processes in the DAN and reorient attention to an exogenous stimulus (Japee et al., 2015; Briggs et al., 2021). Furthermore, the right MFG actively engages when reorienting to distinctive signals from unexpected locations (Carter et al., 2006).

Our findings revealed reduced functional connectivity between the dorsolateral SFG and the right MFG. This change could lead to disruption between VAN and the DAN, which is influential for attention orientation to novel stimuli. This conclusion agrees with the typical description from the tinnitus patients that they unconsciously perceive their tinnitus to be more prominent in quieter situations, regardless of tinnitus duration (Xu et al., 2019).

In addition, we also discovered that subjects with tinnitus developed within six months (ROT group) demonstrated statistically higher functional connectivity than subjects with persistent tinnitus (PT group). Persistent tinnitus subjects (PT) also demonstrated higher THQ, SAS, and SDS scores than the recent-onset subjects (ROT). This result indicated that tinnitus patients would experience more difficulties reorienting their attention away from tinnitus once it lasted longer than six months (from recent-onset to persistent).

### 4.5 Reduced Functional Connectivity between Bilateral Dorsolateral Superior Frontal Gyrus (SFG) and Superior Parietal Gyrus (SPG) in Both Recent-Onset and Persistent Tinnitus Subjects

The main functions of the SFG comprise spontaneous attention regulation and top-down processing. Existing literature suggested that the SPG became more active during a task-free resting state. Since the SFG acts as a critical component of the superior parietal lobule (SPL), the SPG demonstrates a strong connection with the occipital lobe and involves somatosensory and visuospatial stimuli integration, written language, and working memory (Berlucchi & Vallar, 2018). Existing literature also reported SPG’s implications in shifting attention between visual targets and spatial-related attention shifts state (Lin et al., 2021). Our findings revealed no significant difference between the functional connectivity level between the ROT and PT group. However, both groups demonstrated a reduced functional connectivity level compared to the healthy control group (CN). Thus, this finding indicated that reduced functional connectivity between the dorsolateral SFG and the SPG could disrupt tinnitus patients’ working memory, regardless of tinnitus duration.

### 4.6 Clinical Significance of Our Findings in Tinnitus Management

Existing literature indicated that the level of tinnitus distress within six months of initial onset predicts the long-term level of tinnitus distress in patients after six months of onset. Patients who perceive higher levels of disruption from tinnitus are more likely to develop persistent and debilitating tinnitus. Multiple findings from our study indicated that patients with recent-onset tinnitus demonstrate the reduced capability of top-down attention and stimuli monitoring and orientations. Therefore, clinicians should provide effective tinnitus management strategies for patients with recent-onset tinnitus (Kleinstäuber and Weise, 2020).

Considering that the cause of tinnitus can be multifactorial, there is no resolute treatment plan for tinnitus. Nevertheless, clinicians can still effectively manage tinnitus with the help of multidisciplinary options. According to the clinical practice guideline for tinnitus from the American Academy of Otolaryngology-Head and Neck Surgery, patient education and counseling, hearing amplification, sound therapy, and cognitive-behavioral therapy (CBT) should be implemented individually or in combination for tinnitus management (Tunkel et al., 2014; Zenner et al., 2016; Liu et al., 2021; Osuji, 2021).

## 5 Conclusion

Our project indicated a reduced activity level within the dorsolateral SFG (left and right) and gyrus rectus, using ALFF and ReHo analyses. Patients with persistent tinnitus demonstrated a higher activity level in the gyrus rectus than patients with recent-onset tinnitus. Furthermore, our follow-up Voxel-Wise functional connectivity revealed decreased connection activity between the dorsolateral SFG (left and right) and right Superior Parietal Gyrus (SPG), right Middle Frontal Gyrus (MFG), and left medial Superior Frontal Gyrus (mSFG) for subjects with recent-onset (ROT) and persistent tinnitus (PT), compared to the healthy control group. Patients with recent-onset tinnitus demonstrate a higher level of functional connectivity than those with persistent tinnitus. Our data suggested that patients with persistent tinnitus are more likely to experience difficulties in external stimuli monitoring and attention reorientation than patients with recent-onset tinnitus. In addition, patients who perceive higher levels of disruption from tinnitus are more likely to develop persistent and debilitating tinnitus. Therefore, we strongly recommend that clinicians implement effective tinnitus management strategies for patients with recent-onset tinnitus as soon as possible.

## Data Availability

All data produced in the present work are contained in the manuscript

## Author Contributions

Haoliang Du^1^: Conceptualization, Methodology, Investigation, Formal analysis, Writing-Original Draft Preparation. **Xu Feng** ^**1†**^: Resources, Investigation, Data Curation, Writing - Review & Editing. **Xiaoyun Qian**^**2**^: Supervision, Project administration, Writing - Review & Editing, Funding acquisition. **Jian Zhang**^**3**^: Resources, Software, Validation, Visualization, Formal Analysis. **Bin Liu**^**4**^: Resources, Software, Validation, Formal Analysis. **Ao Li**^**5**^: Funding Support, Resources, Validation. **Xia Gao*:** Supervision, Project administration, Funding acquisition. **Zhichun Huang*:** Supervision, Project administration, Funding acquisition

All authors contributed to the article and approved the submitted version.

## Acknowledgments

We would like to express our most sincere appreciation to Dr. Xia Gao and Dr. Zhichun Huang for their supervision, guidance, and encouragement throughout this project. We would also like to extend our gratitude to the Department of Otolaryngology-Head and Neck Surgery and the Department of Radiology at Nanjing Zhongda Hospital, affiliated with Southeast University, for graciously sharing precious clinical data with us. All authors are supported by The Project of Invigorating Health Care through Science, Technology, and Education.

