## Supplementary material for "Recent-Onset and Persistent Tinnitus: Uncover the Differences in Brain Activities using Resting-State Functional Magnetic Resonance Imaging Technologies": Figures

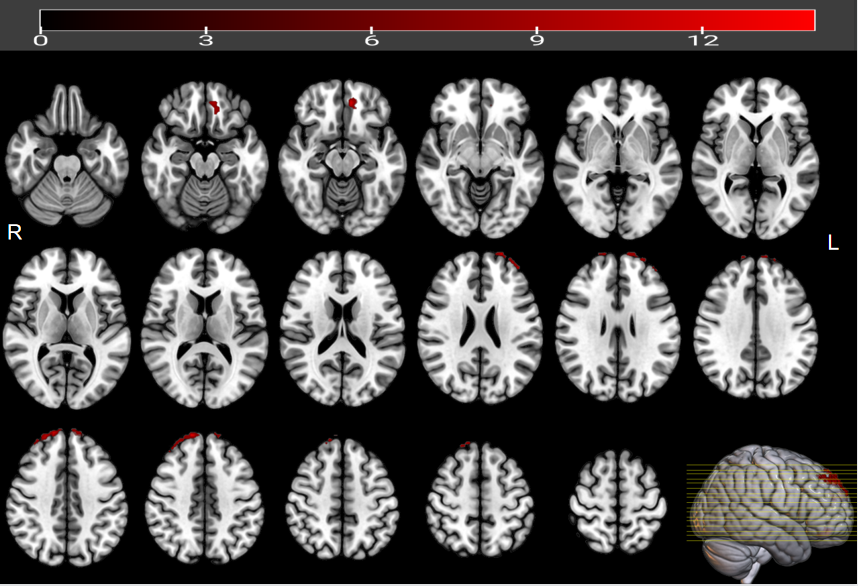
**Fig. 1.** Significant ALFF value differences in the left and right dorsolateral SFG and Rectus Gyrus for both Recent-onset Tinnitus (ROT) and Persistent Tinnitus (PT) groups, compared to the healthy control group (CN).

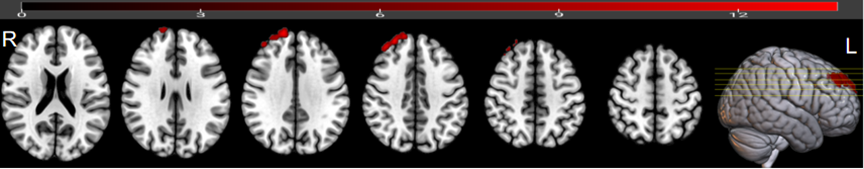
**Fig. 2.** Significant ReHo value differences in the right dorsolateral SFG for Recent-onset Tinnitus (ROT) and Persistent Tinnitus (PT) groups, compared to the CN group.

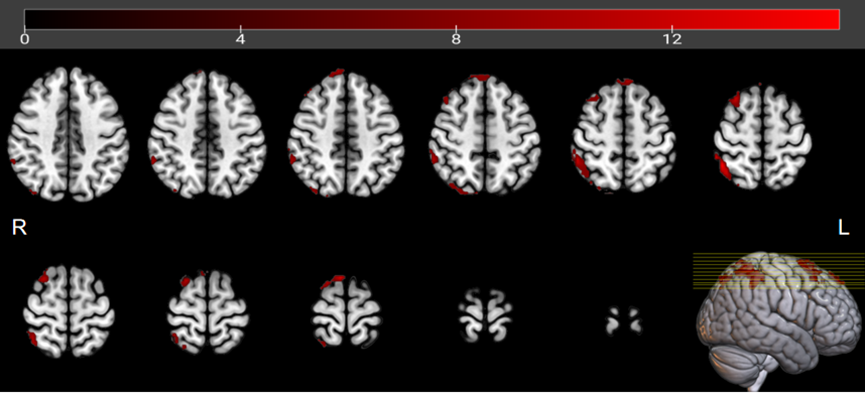
**Fig. 3** Significant FC patterns between the ROIs (dorsolateral SFG, left) and the right SFG, left medial Superior Frontal Gyrus (SFG), and right Superior Parietal Gyrus (SPG).

**Fig. 4.** Significant FC patterns between the right dorsolateral SFG and the right Middle Frontal Gyrus (MFG), right medial Superior Frontal Gyrus (SFG), and right Superior Parietal Gyrus (SPG) in Recent-onset Tinnitus (ROT) and Persistent Tinnitus (PT) groups, compared to the CN group.

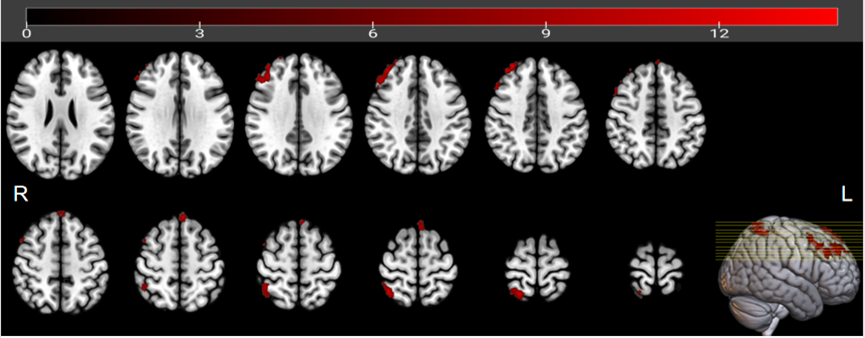

**Fig. 5.** Significant FC patterns between the right dorsolateral SFG and right Middle Frontal Gyrus (MFG), right Superior Parietal Gyrus (SPG), and right dorsolateral SFG in Recent-onset Tinnitus (ROT) and Persistent Tinnitus (PT) groups, compared to the CN group.

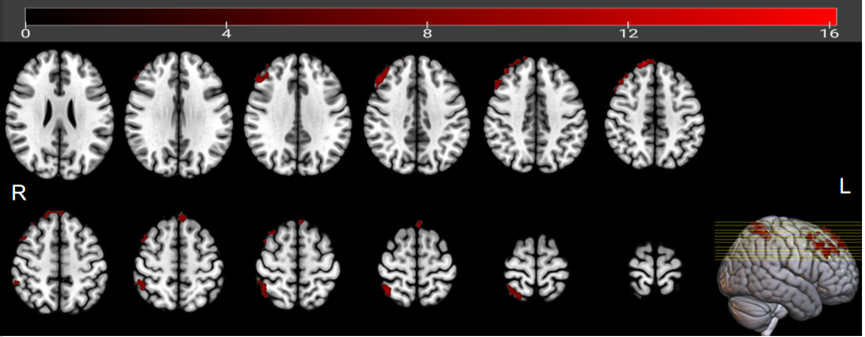
