## Supplementary material for "Recent-Onset and Persistent Tinnitus: Uncover the Differences in Brain Activities using Resting-State Functional Magnetic Resonance Imaging Technologies": Table

Tables

**Table 1**

Subject Characteristics of the Recent-onset Tinnitus Group (ROT), Persistent Tinnitus Group (PT), and Control Group (CN)

|  | ROT Group | PT Group | CN Group | p-value |
| --- | --- | --- | --- | --- |
| Age (Year) | 45.32±2.93 | 42.68±2.33 | 37.38±1.84 | >0.05 |
| Gender (Male: Female) | 14:11 | 14:14 | 20:9 | >0.05 |
| Education Duration (Year) | 9.83±2.11 | 9.22±1.96 | 10.12±2.43 | >0.05 |
| THQ Total Score | 40.67±3.89 | 44.97±4.27 | -- | <0.05 |
| SAS Score | 35.12±1.07 | 37.57±1.51 | -- | <0.05 |
| SDS Score | 37.96±1.85 | 39.06±2.12 | -- | <0.05 |

Data are represented as Mean ± SD

**Table 2a** Decreased ALFF activities in both Recent-onset Tinnitus (ROT) and Persistent Tinnitus (PT) with than in the control group (CN)

| Cluster number | Cluster size (Voxel) | Peak MNI coordinate | | | Peak MNI  coordinate region | F Value | T Value Difference between ROT and PT | T Value Difference between ROT and CN | T Value Difference between PT and CN |
| --- | --- | --- | --- | --- | --- | --- | --- | --- | --- |
|  |  | X | Y | Z |  |  |  |  |  |
| 1 | 25 | -12 | 45 | -15 | Left Gyrus Rectus | 14.36 | -2.93 | -5.18 | -2.25 |
| 2 | 49 | -15 | 54 | 42 | Left Dorsolateral SFG | 15.79 | No significant difference | -4.60 | -4.34 |
| 3 | 73 | 15 | 54 | 45 | Right Dorsolateral SFG | 13.85 | No significant difference | -3.86 | -4.26 |

**Table 2b** Decreased ReHo activities in both Recent-onset Tinnitus (ROT) and Persistent Tinnitus (PT) with than in the control group (CN)

| Cluster number | Cluster size（voxels） | Peak MNI coordinate | | | Peak MNI coordinate region | F Value | T Value Difference between ROT and PT | T Value Difference between ROT and CN | T Value Difference between PT and CN |
| --- | --- | --- | --- | --- | --- | --- | --- | --- | --- |
|  |  | X | Y | Z |  |  |  |  |  |
| 1 | 160 | 39 | 45 | 39 | Right Dorsolateral SFG | 10.02 | No significant difference | -4.06 | -4.77 |

**Table 3a** Decreased activities in Voxel-Wise Functional Connectivity (FC) ALFF cluster 2 for both Recent-onset Tinnitus (ROT) and Persistent Tinnitus (PT) groups than in the control group (CN)

| Cluster number | Cluster size（voxels） | Peak MNI coordinate | | | Peak MNI coordinate region | F Value | T Value Difference between ROT and PT | T Value Difference between ROT and CN | T Value Difference between PT and CN |
| --- | --- | --- | --- | --- | --- | --- | --- | --- | --- |
|  |  | X | Y | Z |  |  |  |  |  |
| 1 | 150 | 45 | -51 | 60 | Right Superior Parietal Gyrus (SPG) | 14.86 | No significant difference | -3.78 | -4.66 |
| 2 | 83 | 6 | 15 | 72 | Right Dorsolateral SFG | 14.98 | No significant difference | -3.62 | -3.84 |
| 3 | 43 | 9 | 54 | 48 | Left Superior Medial Frontal Gyrus | 13.44 | No significant difference | -3.36 | -4.23 |

**Table 3b** Decreased activities in Voxel-Wise Functional Connectivity (FC) ALFF cluster 3 for both Recent-onset Tinnitus (ROT) and Persistent Tinnitus (PT) groups than in the control group (CN)

| Cluster number | Cluster size（voxels） | Peak MNI coordinate | | | Peak MNI coordinate region | F Value | T Value Difference between ROT and PT | T Value Difference between ROT and CN | T Value Difference between PT and CN |
| --- | --- | --- | --- | --- | --- | --- | --- | --- | --- |
|  |  | X | Y | Z |  |  |  |  |  |
| 1 | 132 | 39 | 42 | 39 | Right Middle Frontal Gyrus (MFG) | 14.86 | 3.18 | -4.58 | -5.09 |
| 2 | 46 | -3 | 42 | 57 | Left Medial Superior Frontal Gyrus | 14.98 | No significant difference | -3.82 | -4.60 |
| 3 | 93 | 36 | -57 | 66 | Right Superior Parietal Gyrus | 13.44 | No significant difference | -4.36 | -4.07 |

**Table 3c** Decreased activities in Voxel-Wise Functional Connectivity (ReHo Cluster 1) for both Recent-onset Tinnitus (ROT) and Persistent Tinnitus (PT) groups than in the control group (CN)

| Cluster number | Cluster size（voxels） | Peak MNI coordinate | | | Peak MNI coordinate region | F Value | T Value Difference between ROT and PT | T Value Difference between ROT and CN | T Value Difference between PT and CN |
| --- | --- | --- | --- | --- | --- | --- | --- | --- | --- |
|  |  | X | Y | Z |  |  |  |  |  |
| 1 | 120 | 48 | 30 | 36 | Right Middle Frontal Gyrus (MFG) | 14.86 | 3.89 | -2.79 | -5.00 |
| 2 | 80 | 15 | 51 | 48 | Right Dorsolateral Superior Frontal Gyrus (SFG) | 14.98 | No significant difference | -4.17 | -4.66 |
| 3 | 96 | 36 | -51 | 66 | Right Superior Parietal Gyrus (SPG) | 13.44 | No significant difference | -3.31 | -4.49 |
